# Large language models to differentiate vasospastic angina using patient information

**DOI:** 10.1101/2023.06.26.23291913

**Authors:** Yuko Kiyohara, Satoshi Kodera, Masaya Sato, Kota Ninomiya, Masataka Sato, Hiroki Shinohara, Norifumi Takeda, Hiroshi Akazawa, Hiroyuki Morita, Issei Komuro

**Author notes:** Corresponding author (SK).

## Abstract

**Background:** Vasospastic angina is sometimes suspected from patients’ medical history. It is essential to appropriately distinguish vasospastic angina from acute coronary syndrome because its standard treatment is pharmacotherapy, not catheter intervention. Large language models have recently been developed and are currently widely accessible. In this study, we aimed to use large language models to distinguish between vasospastic angina and acute coronary syndrome from patient information and compare the accuracies of these models.

**Method:** We searched for cases of vasospastic angina and acute coronary syndrome which were written in Japanese and published in online-accessible abstracts and journals, and randomly selected 66 cases as a test dataset. In addition, we selected another ten cases as data for few-shot learning. We used generative pre-trained transformer-3.5 and 4, and Bard, with zero- and few-shot learning. We evaluated the accuracies of the models using the test dataset.

**Results:** Generative pre-trained transformer-3.5 with zero-shot learning achieved an accuracy of 52%, sensitivity of 68%, and specificity of 29%; with few-shot learning, it achieved an accuracy of 52%, sensitivity of 26%, and specificity of 86%. Generative pre-trained transformer-4 with zero-shot learning achieved an accuracy of 58%, sensitivity of 29%, and specificity of 96%; with few-shot learning, it achieved an accuracy of 61%, sensitivity of 63%, and specificity of 57%. Bard with zero-shot learning achieved an accuracy of 47%, sensitivity of 16%, and specificity of 89%; with few-shot learning, this model could not be assessed because it failed to produce output.

**Conclusion:** Generative pre-trained transformer-4 with few-shot learning was the best of all the models. The accuracies of models with zero- and few-shot learning were almost the same. In the future, models could be made more accurate by combining text data with other modalities.

## Introduction

Vasospastic angina (VSA) is characterized by transient constriction of the coronary artery, causing angina and other nonspecific symptoms [1–3]. VSA is known to be caused by various factors, including emotional stress, exposure to low temperatures, and resting during the period from midnight to early morning [3–7]. Conversely, acute coronary syndrome (ACS), which is mostly caused by diseases that differ from VSA, frequently presents with chest pain during physical exertion following a prolonged history of smoking, dyslipidemia, hypertension, or diabetes mellitus [8–10]. Because VSA shares some symptoms with ACS, which requires urgent catheter intervention treatment, distinguishing between VSA and ACS is often challenging at the screening stage [3,4,8–11]. In cases where it is difficult to distinguish between the two, VSA is confirmed by coronary spasm provocation testing, as described in the guideline and international consensus [12,13]. However, provocation testing is an invasive procedure that carries the risk of serious complications [14]. Therefore, there is a need for a noninvasive method of screening for VSA in patients with angina.

In recent years, large language models (LLMs) have demonstrated great potential in a broad range of fields including medicine [15–17]. In the medical field, generative pre-trained transformer 4 (GPT-4) has achieved high scores in medical licensing examinations in the United States and Japan [18–20]. In real-world clinical settings, physicians rely on patient information gathered through interviews and examinations. However, to the best of our knowledge, there has been no research on whether LLMs can effectively use patient information to distinguish between specific diseases. Furthermore, although GPT-4 has multilingual capabilities, much of the research conducted using this model has focused on English documents [19,21]. In this study, we aimed to use LLMs to distinguish between VSA and ACS from patient information written in Japanese and compare the accuracies of the models.

## Materials and methods

An overview of this study is shown in Fig 1.

**Fig 1.**
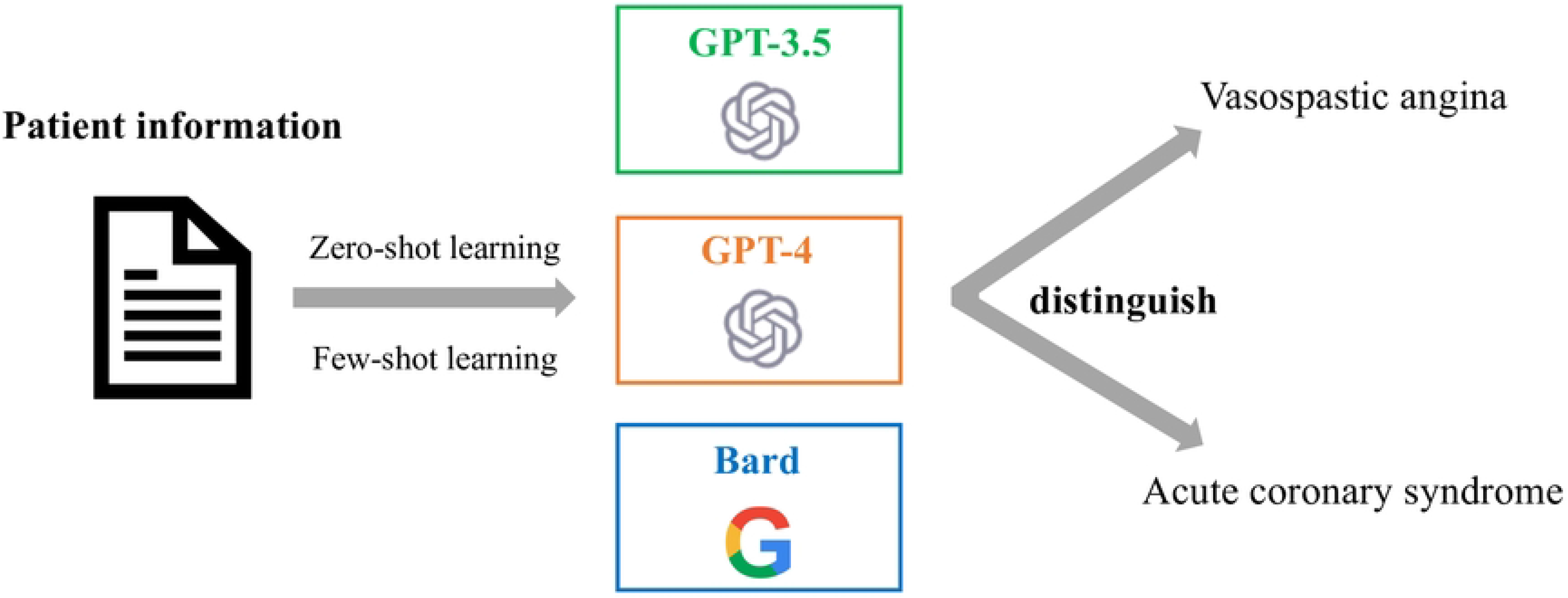
The flow of this study. The overview of this study is shown.

### Case selection

In a previous study, we continuously collected cases of VSA and ACS published in abstracts of the Japanese Society of Internal Medicine regional conferences and open-access journals. In the present study, we randomly selected 66 of these cases (38 VSA and 28 ACS cases) as a test dataset [22]. We defined ACS to include both unstable angina and acute myocardial infarction. In addition, we selected another ten cases (five VSA and five ACS cases) from the previously collected cases as data for few-shot learning. Because the data used in this study were publicly accessible, there were no ethical concerns. Nonetheless, we handled cases following the principles outlined in the Declaration of Helsinki.

### Data extraction

We extracted data comprising age, sex, medical history, past medical history, and medication from the selected abstracts and case reports, and organized them accordingly (S1 Fig). Medication doses were not extracted, and the English notation of medication was translated to Japanese and standardized.

### GPT-3.5 and GPT-4

We used ChatGPT Plus (OpenAI), which is based on generative pre-trained transformer-3.5 (GPT-3.5) and GPT-4 [20]. In addition, we adopted the zero- and few-shot learning approaches. To select appropriate cases to include in the data for few-shot learning, YK and SK discussed and identified typical cases of VSA and ACS. We selected ten representative cases from the data that were left behind after selecting the test dataset, and these cases were used as data for few-shot learning.

At the beginning of the prompts, we asked GPT-3.5 and GPT-4 to answer whether each case was predicted to be VSA or any other coronary artery disease. Technically, ACS can be caused also by VSA, although rare, and therefore, we used the phrase of “any other coronary artery disease” rather than ACS. In the zero-shot learning tests, we input the cases from the test dataset just after the above question. In the few-shot learning tests, we inserted the set comprising each case and its correct answer, for the ten cases of the learning data (S2 Fig). We performed the experiment from May 10 to May 20, 2023. We used a new session for each case by clearing all conversations before inputting any prompts.

### Bard

We used Bard (Google) because it was announced on May 11, 2023 that Bard could respond to Japanese text. We input the same request as we input to GPT-3.5 and GPT-4. In the zero-shot learning tests, we input the cases in the test dataset following the request sentence. In the few-shot learning tests, we input the same set of ten cases (with their correct answers) as we input to GPT-3.5 and GPT-4. We performed the experiment from May 22 to May 24, 2023. We reset the conversations every time we input a prompt.

### Evaluation of model accuracy

In each of the three groups of experiments (GPT-3.5, GPT-4, and Bard), we compared their answers with the correct answers, and calculated accuracy, sensitivity, specificity, precision, and F-score. We compared the accuracies achieved by the three LLMs. The threshold for statistical significance was set at 0.05. The accuracy of each model was calculated with a 95% confidence interval (CI) using the binomial test, implemented in R (R Foundation for Statistical Computing, Vienna, Austria).

### Comparison with cardiologists’ accuracy and accuracy of each model

In the previous study, we reported the accuracy with which cardiologists answered the cases in the test dataset [22]. In this study, for each case, we evaluated the answer given by each model with reference to those given by the cardiologists.

### Sensitivity analysis

As a sensitivity analysis, we selected the subset of the test dataset comprising the cases for which more than half of the cardiologists answered correctly in the previous study [22]. In 45 out of 66 cases, more than 50% of the cardiologists answered correctly. We hypothesized that these cases contain enough information to distinguish between VSA and ACS. Therefore, in the sensitivity analysis, we used these 45 cases only.

## Results

### Accuracy evaluation of GPT-3.5, GPT-4, and Bard

Table 1 shows the accuracy achieved in the three groups of experiments. For GPT-3.5 with zero-shot learning, the accuracy was 52% (95% CI: 39–64%), sensitivity was 68%, specificity was 29%, precision was 57%, and F-score was 62%; with few-shot learning, the accuracy was 52% (95% CI: 39–64%), sensitivity was 26%, specificity was 86%, precision was 71%, and F-score was 39%. For GPT-4 with zero-shot learning, the accuracy was 58% (95% CI: 45–70%), sensitivity was 29%, specificity was 96%, precision was 92%, and F-score was 44%; with few-shot learning, the accuracy was 61% (95% CI: 48–72%), sensitivity was 63%, specificity was 57%, precision was 67%, and F-score was 65%. For Bard with zero-shot learning, the accuracy was 47% (95% CI: 35–60%), sensitivity was 16%, specificity was 89%, precision was 67%, and F-score was 26%; with few-shot learning, Bard failed to respond to the input data.

**Table 1.** The main results of accuracies of the models.

|  | GPT-3.5/zero | GPT-3.5/few | GPT-4/zero | GPT-4/few | Bard/zero | Bard/few |
| --- | --- | --- | --- | --- | --- | --- |
| Accuracy | 51.5<br>(95%CI: 38.9 - 64.0) | 51.5<br>(95%CI: 38.9 - 64.0) | 57.6<br>(95%CI: 44.8 - 69.7) | 60.6<br>(95%CI: 47.8 - 72.4) | 47.0<br>(95%CI: 34.6 - 59.7) | NA |
| Sensitivity | 68.4 | 26.3 | 28.9 | 63.2 | 15.8 | NA |
| Spesificity | 28.6 | 85.7 | 96.4 | 57.1 | 89.3 | NA |
| Precision | 56.5 | 71.4 | 91.7 | 66.7 | 66.7 | NA |
| F-score | 61.9 | 38.5 | 44.0 | 64.9 | 25.5 | NA |

The accuracy achieved in each of the three AI models (GPT-3.5, GPT-4, and Bard) with zero- and few-shot learning is shown.

### Comparison with cardiologists’ accuracy and result of each model

Fig 2 shows the results of GPT-4 with few-shot learning with reference to the cardiologists’ accuracy (the proportion of cardiologists who answered correctly). GPT-4 tended to correctly answer cases for which the cardiologists’ accuracy was high. This tendency was observed for the other models except for GPT-3.5 with zero-shot learning (S3–6 Figs).

**Fig 2.**
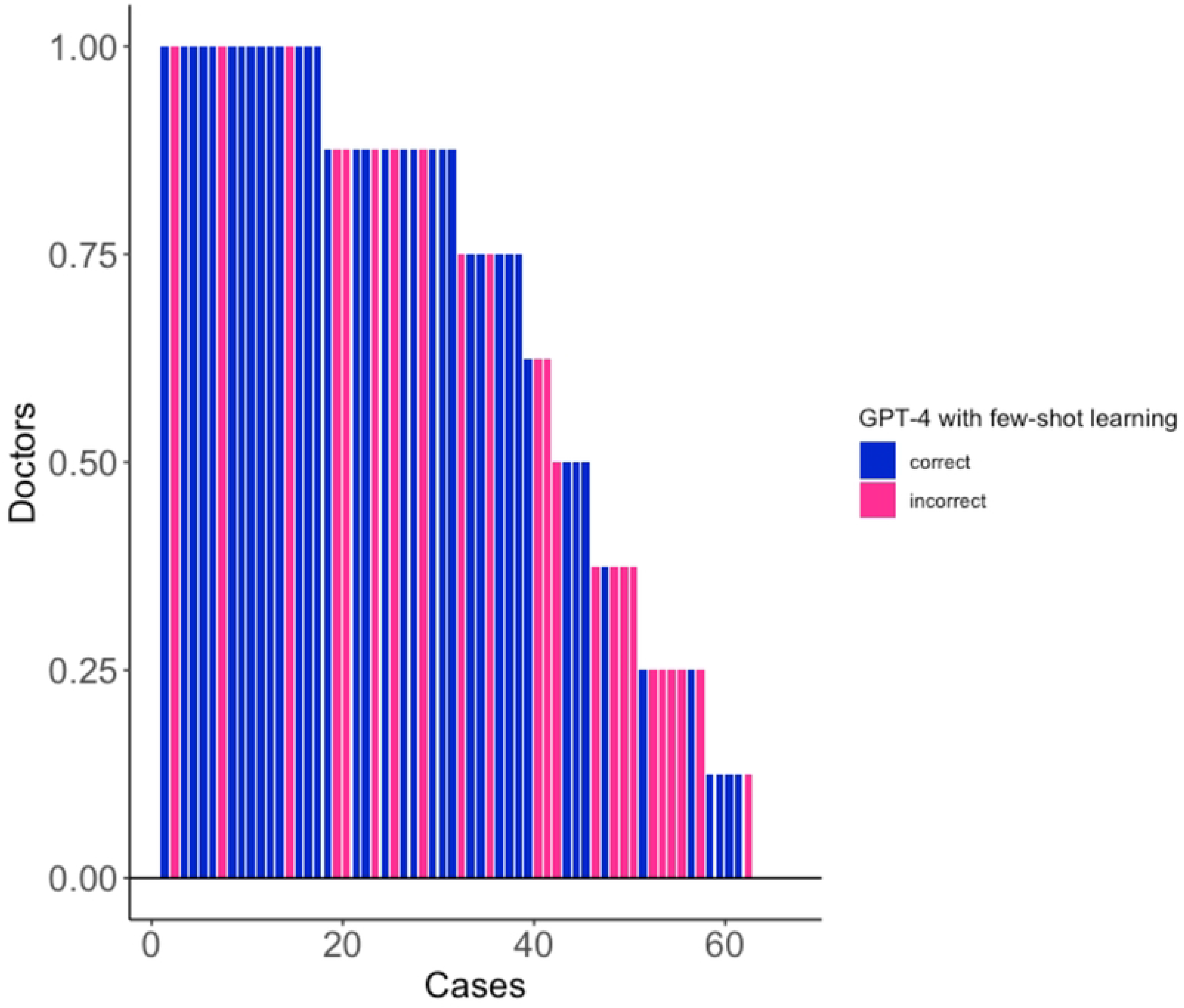
Comparison with cardiologists’ accuracy and GPT-4 with few-shot learning. The result of GPT-4 with few-shot learning is shown with reference to the proportion of cardiologists who answered correctly.

### Sensitivity analysis

In the sensitivity analysis, the results were almost the same as the main results (Table 2). GPT-4 yielded the highest accuracy of the three models. In GPT-4 with zero-shot learning, the accuracy was 76%, sensitivity was 50%, specificity was 100%, precision was 100%, and F-score was 67%; with few-shot learning, the accuracy was 71%, sensitivity was 77%, specificity was 65%, precision was 68%, and F-score was 72%.

**Table 2.**
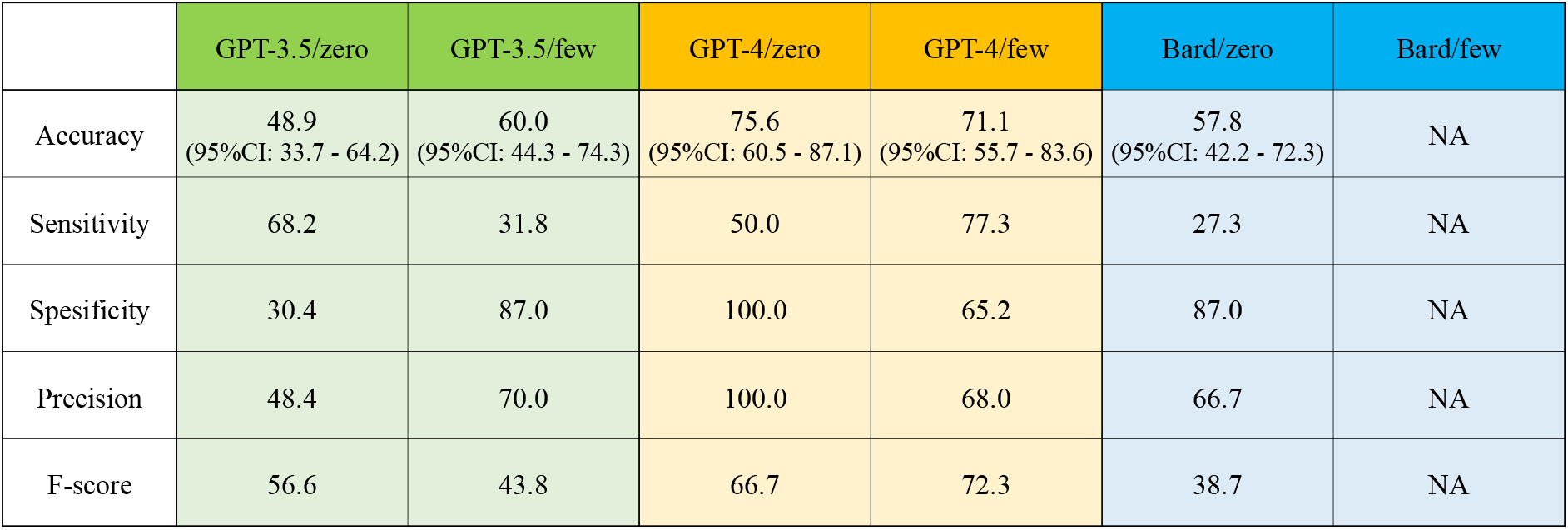
The results of the sensitivity analysis.

|  | GPT-3.5/zero | GPT-3.5/few | GPT-4/zero | GPT-4/few | Bard/zero | Bard/few |
| --- | --- | --- | --- | --- | --- | --- |
| Accuracy | 48.9<br>(95%CI: 33.7 - 64.2) | 60.0<br>(95%CI: 44.3 - 74.3) | 75.6<br>(95%CI: 60.5 - 87.1) | 71.1<br>(95%CI: 55.7 - 83.6) | 57.8<br>(95%CI: 42.2 - 72.3) | NA |
| Sensitivity | 68.2 | 31.8 | 50.0 | 77.3 | 27.3 | NA |
| Spesificity | 30.4 | 87.0 | 100.0 | 65.2 | 87.0 | NA |
| Precision | 48.4 | 70.0 | 100.0 | 68.0 | 66.7 | NA |
| F-score | 56.6 | 43.8 | 66.7 | 72.3 | 38.7 | NA |

The results of experiments for the subset of the test dataset for which more than half of the cardiologists answered correctly are shown.

## Discussion

In this study, we used GPT-3.5, GPT-4, and Bard with zero- and few-shot learning for the purpose of distinguishing between VSA and ACS from patient information. The results of the study can be summarized as follows: 1) GPT-4 with few-shot learning yielded the most accurate results of the three LLMs, 2) there were no significant differences in accuracy between zero- and few-shot learning, and 3) our study was unique in that it processed data in the form of Japanese text instead of English text.

GPT-4 was able to distinguish VSA and ACS from patient information and its accuracy was almost the same as that of medical students. In a previous study, we compared a variant model of BERT (Google) with cardiologists and medical students [22,23]. The test data were the same as those used in this study. For cardiologists, the accuracy was 68%, sensitivity was 58%, and specificity was 82%, whereas for medical students, the accuracy was 61%, sensitivity was 40%, and specificity was 89% [22]. In the current study, GPT-4 with few-shot learning achieved almost the same accuracy as medical students. Because we collected cases from conference abstracts of oral presentations and peer-reviewed articles, which ensured linguistic consistency and no unique abbreviations or variations in sentences, we certainly used high-quality data. However, the performance of GPT-4 in distinguishing VSA from ACS did not match that of cardiologists. This implies that the advantages of enhancing the quality of data are limited. Improvements to the methodology of models are required to obtain further improvements in performance.

Surprisingly, we found no significant differences in accuracy between zero- and few-shot learning. Previously, few-shot learning was reported to increase the accuracy of LLMs [24]. However, although we used GPT-3.5 and GPT-4 with both zero- and few-shot learning, there were almost no differences in accuracy between the two methods. This suggests that few-shot learning may not necessarily improve accuracy. Fine-tuning is another promising method for enhancing the accuracy of models. Fine-tuning involves adjusting the parameters and architecture of models, using training data to adapt them to specific tasks [25,26]. Initially, we attempted to conduct fine-tuning using the remaining data for the purpose of tailoring the models to our specific tasks. However, applying fine-tuning to GPT-4 was impossible as of May 20, 2023. In the future, fine-tuning could potentially improve the accuracy of GPT-4 even further.

Our study has the potential to contribute to the advancement of natural language processing of Japanese medical data. A comprehensive review showed that almost 90% of the publications about clinical natural language processing addressed the processing of English text data, whereas only 0.62% were on Japanese text data [27]. The multilingual capability of GPT-4 will facilitate research on Japanese text. Much more research specific to the Japanese population is required because there are some diseases, such as VSA, which are more common in Japan than elsewhere.

There is scope for improvement in the accuracy of artificial intelligence (AI) models and their applications to other diseases. It is expected that the accuracy of AI models can be improved by combining other types of data with text data. In a previous study, an AI model was developed that used numerical and image data, including laboratory data and resting 12-lead electrocardiograms, to determine the presence or absence of coronary abnormalities in patients with chest pain [28]. “Generalist” medical AI, capable of flexibly incorporating data modalities including text data, numerical data (such as examination findings), and image data (such as electrocardiograms), could potentially be useful in clinical settings [29]. Furthermore, the methodology of this study can be applied to diseases other than VSA, thereby facilitating the development of AI models to distinguish between various diseases. There are some limitations to this study. First, the number of conference abstracts and published papers collected in the study was limited, and it is difficult to generalize its results. One possible solution to the scarcity of data is the extraction of data from electronic health records [30]. Although electronic health records contain patient information, the quality of this information may be inadequate. Electronic health records are often written by busy physicians and nurses; they therefore contain many abbreviations and incorrect or vague expressions, and often ignore grammar [31,32]. Therefore, the quality of electronic health record data is considered to be lower than that of the abstracts and reports used in this study. Second, regarding the content of the data, the fact that the cardiologists’ accuracy was only around 70% suggests that the data used in this study may not have contained sufficient information for distinguishing VSA. That said, in actual clinical practice, it is often difficult to determine VSA solely from patients’ medical history; therefore, the data used in this study could not be necessarily reflective of real-world clinical practice. Third, we asked the models to answer whether each case was VSA or any other coronary artery disease. We believe that this prompt was very effective in distinguishing between VSA and ACS. However, we would like to further improve models by directly comparing VSA and ACS. Fourth, in the data collection process, we defined the ACS group as cases described as “unstable angina” or “acute myocardial infarction.” Cases described using synonyms or different expressions, such as “acute coronary syndrome” or “ACS,” may have been omitted, possibly limiting the quantity of data available for training the AI model and evaluating its accuracy. Future research should aim to collect cases comprehensively, including those described using synonyms and different expressions.

In conclusion, we used GPT-3.5, GPT-4, and Bard to distinguish between VSA and ACS from patient information. The predictive accuracy of GPT-4 with few-shot learning was the best of the three models. In the future, by creating multimodal AI models that combine text data with numerical data, image data, and other types of data, it is expected that the accuracy will be further improved.

## Data Availability

All relevant data are within the manuscript and its Supporting Information files.

## Acknowledgments

We employed GPT-4 to generate initial drafts, which were then critically reviewed. We take full responsibility for the content presented in this article. We thank Edanz (https://jp.edanz.com/ac) for editing a draft of this manuscript.

## Supporting information

**S1 Fig. An example of the data.**

This example was extracted from the following case report published in an open-access journal. We used only the original sentences written in Japanese above in the box. In the figure, we translated the whole text into English to make it easy to understand.

**S2 Fig. The detailed data for few-shot learning.**

This figure shows the ten cases of the learning data used for few-shot learning. In the prompts, we put only the original sentences written in Japanese in the above box. In the figure, we translated the whole text into English to make it easy to understand.

**S3 Fig. Comparison with cardiologists’ accuracy and GPT-3.5 with zero-shot learning.**

The result of GPT-3.5 with zero-shot learning is shown with reference to the proportion of cardiologists who answered correctly.

**S4 Fig. Comparison with cardiologists’ accuracy and GPT-3.5 with few-shot learning.**

The result of GPT-3.5 with few-shot learning is shown with reference to the proportion of cardiologists who answered correctly.

**S5 Fig. Comparison with cardiologists’ accuracy and GPT-4 with zero-shot learning.**

The result of GPT-4 with zero-shot learning is shown with reference to the proportion of cardiologists who answered correctly.

**S6 Fig. Comparison with cardiologists’ accuracy and Bard with zero-shot learning.**

The result of Bard with zero-shot learning is shown with reference to the proportion of cardiologists who answered correctly.

## Notes

### Competing Interest Statement

The authors have declared no competing interest.

### Funding Statement

This work was supported by JSPS KAKENHI (grant no. JP 23H03491).

